# Transcranial magnetic stimulation language mapping analysis revisited: Machine learning classification of 90 patients reveals distinct reorganization pattern in aphasic patients

**DOI:** 10.1101/2020.02.06.20020693

**Authors:** Ziqian Wang, Lucius Fekonja, Felix Dreyer, Peter Vajkoczy, Thomas Picht

## Abstract

Repetitive TMS (rTMS) allows to non-invasively and transiently disrupt local neuronal functioning. Its potential for mapping of language function is currently explored. Given the inter-individual heterogeneity of tumor impact on the language network and resulting rTMS derived functional mapping, we propose to use machine learning strategies to classify potential patterns of functional reorganization. We retrospectively included 90 patients with left perisylvian glioma tumors, world health organization (WHO) grade II-IV, affecting the language network. All patients underwent navigated rTMS language mappings. The severity of aphasia was assessed preoperatively using the Berlin Aphasia Score (BAS), which is adapted to the Aachener Aphasia Test (AAT). After spatial normalization to MNI 152 of all rTMS spots, we calculated the error rate (ER) in each cortical area by automated anatomical labeling parcellation (AAL) and used support vector machine (SVM) as a classifier for significant areas in relation to aphasia. 29 of 90 (32.2%) patients suffered from aphasia. Univariate analysis revealed 11 perisylvian AVOIs’ ERs (eight left, three right hemispheric) that were significantly higher in the aphasic than non-aphasic group (p < 0.05), depicting a broad, bihemispheric language network. After feeding the significant AVOIs into the SVM model, it showed that additional to age (w = 2.95), the ERs of right Frontal_Inf_Tri (w = 2.06) and left SupraMarginal (w = 2.05) and Parietal_Inf (w= 1.80) contributed more than other features to the model. The model’s sensitivity was 89.7%, the specificity was 82.0%, the overall accuracy was 81.1% and AUC was 88.7%. Our results demonstrate an increased vulnerability of the right inferior frontal gyrus to rTMS in patients suffering from aphasia due to left perisylvian gliomas. This confirms a functional relevant involvement of the right frontal area in relation to aphasia. While age as a feature improved our SVM model the most, the tumor location feature didn’t affect the SVM model. This finding indicates that general tumor induced network disconnection is relevant to aphasia and not necessarily related to specific lesion locations. Additionally, our results emphasize the decreasing potential for neuroplasticity with age.

## 1. Introduction

With the paradigm shift from localizationism to associationist and connectomic concepts of function, the challenge to map individual language networks increased. While intraoperative direct electrical stimulation (DES) still defines the gold standard for functional mapping, many efforts are spent on developing non-invasive methods to individually locate eloquent areas for surgical planning and perform safe neurosurgery in patients not undergoing awake procedures.

The development of functional neuroimaging methods improved pre‐operative neurosurgical planning in eloquent brain areas during the last decade.(Duffau, 2015) These developments consist of noninvasive imaging techniques such as Magnetoencephalography (MEG), Positron emission tomography (PET) and functional magnetic resonance imaging (fMRI), respectively. (Cohen, 1968, 1972) (Ter-Pogossian *et al*., 1975) (Kwong *et al*., 1992; Ogawa *et al*., 1992) In 1985, Barker proposed the first magnetic stimulator designed to stimulate the human brain transcranially.(Barker *et al*., 1985) Later, repetitive TMS (rTMS) became a powerful TMS modality, capable of regionally blocking or enabling cortical processes.(Wassermann, 1998) With the capability to modify the neuronal activity of local and distant cortical areas by series or trains of pulses, rTMS became another non-invasive tool for clinical treatments and cortical functional mapping with the potential of neural network modeling.(Wassermann and Lisanby, 2001) Nevertheless, it’s clinical reliability and usefulness for cognitive mapping remains debatable to this day.(Schwarzer *et al*., 2018)

Language function consists of a large, distributed network.(Duffau *et al*., 2014) The nature of this network poses a major challenge to rTMS Cortical language areas seem to be volatile, with very few, if any, indispensable functional regions associated with the peritumoral cortex, its dynamic processes and its neuroplasticity..(Duffau *et al*., 2003; Duffau, 2015) The interlinked brain regions encoding function within large-scale networks are currently simplified and described as associationist and hodotopical models of connectivity.(Catani *et al*., 2012; Duffau *et al*., 2014) Tumor induced neural-reorganization mechanisms further increase the challenge to map language in general. According to Duffau’s and Mesulams hodotopical models of connectivity, the central nervous system (CNS) is organized in parallel networks that not only interact but also can compensate each other to a certain extent.(Mesulam, 1990; Duffau, 2014, 2015) Neuroplasticity is a dynamic process that enables redistribution within remote networks and plays a central part in ontogeny, learning as well as recovery after brain injury.(Duffau, 2005, 2015) Cognitive mapping is additionally complicated by the non-specificity of tasks resulting in uncontrolled co-activations and by the patient’s capability and willingness to cooperate.

To understand how the brain’s structural architecture supports and enables cognitive processes and further advance the current understanding of brain network, researchers are developing computational models and graph-based analysis to investigate the function of brain networks, for example to determine how neuronal activity propagates along structural connections.(Lynn and Bassett, 2019) (Aerts *et al*., 2018; Hahn *et al*., 2019) Current models of language function emphasize above mentioned perisylvian and hodotopic distribution and consist of distributed cortical circuits.

For these reasons, it is necessary to critically question the performance of rTMS for mapping language function in order to better understand its potential and identify suitable use cases. In this study, we describe and investigate the clinical course of 90 patients with brain tumors in the language area and their rTMS-based language maps using spatial normalization methods and a machine learning support vector machine (SVM) model to generate data-driven classification.

## 2. Methods

### 2.1 Participants

We retrospectively included 90 patients (41 female, 49 male., mean age 48.86±14.12, age range 21-82) of a prospectively collected cohort of patients with left perisylvian glioma world health organization (WHO) grade II(12), III(42) and IV(36), affecting the human language network who were preoperatively examined with rTMS (Table 1).(Fekonja *et al*., 2019) Handedness was determined using the Edinburgh handedness inventory.(Oldfield, 1971) The exclusion criteria were: 1. Frequent generalized seizures (more than one per week); 2. Aphasia with more than 28% errors in the baseline object naming task and 3. multiple neoplasms. All patients gave their informed written consent, in line with the declaration of Helsinki as well as approved by the local Human Research Ethics Committee.

**Table 1.**
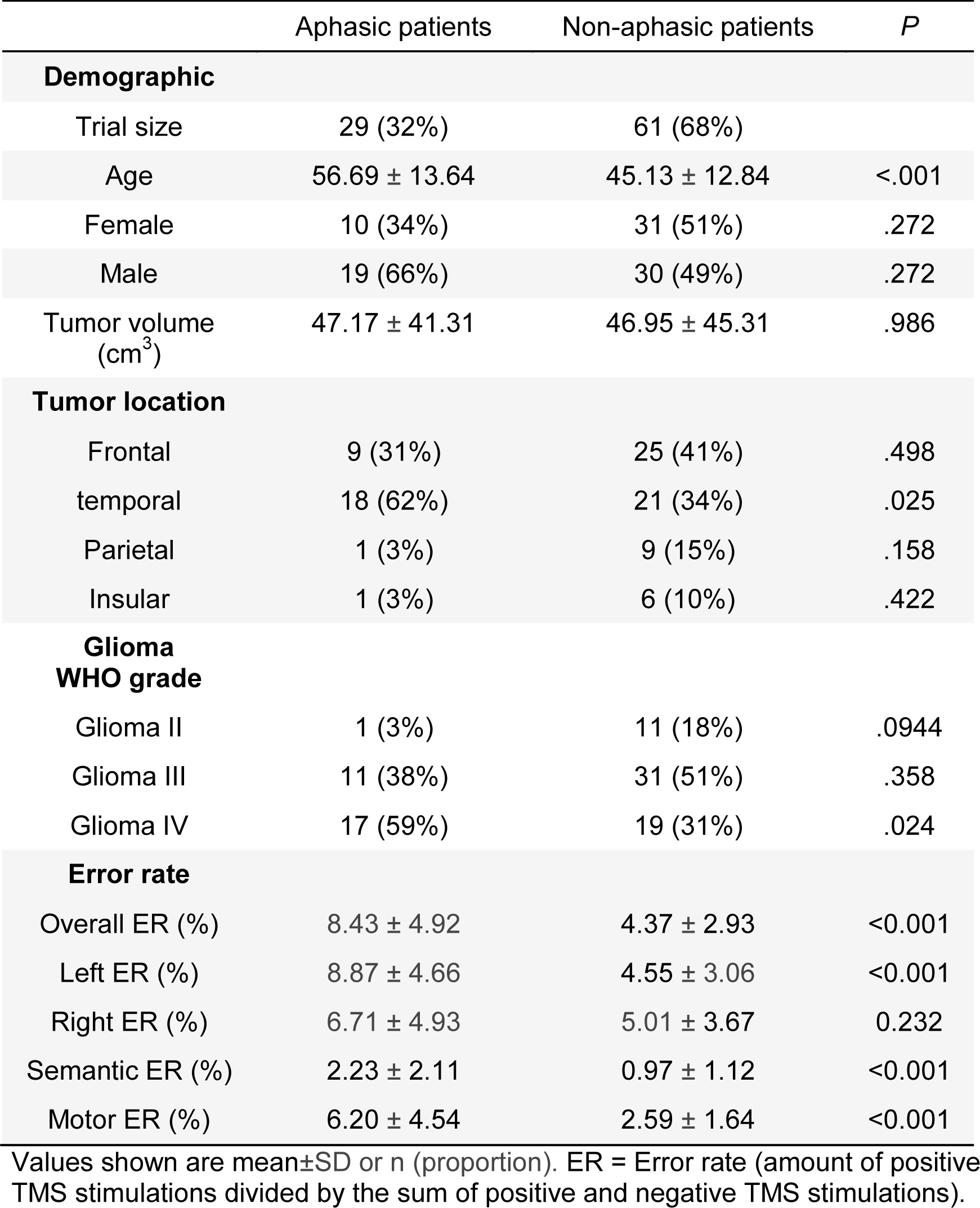
Demographic and neuropathological overview of patient cohort.

|  | Aphasic patients | Non-aphasic patients | *P* |
| --- | --- | --- | --- |
| **Demographic** |  |  |  |
| Trial size | 29 (32%) | 61 (68%) |  |
| Age | 56.69 ± 13.64 | 45.13 ± 12.84 | <.001 |
| Female | 10 (34%) | 31 (51%) | .272 |
| Male | 19 (66%) | 30 (49%) | .272 |
| Tumor volume (cm<sup>3</sup>) | 47.17 ± 41.31 | 46.95 ± 45.31 | .986 |
| **Tumor location** |  |  |  |
| Frontal | 9 (31%) | 25 (41%) | .498 |
| temporal | 18 (62%) | 21 (34%) | .025 |
| Parietal | 1 (3%) | 9 (15%) | .158 |
| Insular | 1 (3%) | 6 (10%) | .422 |
| **Glioma WHO grade** |  |  |  |
| Glioma II | 1 (3%) | 11 (18%) | .0944 |
| Glioma III | 11 (38%) | 31 (51%) | .358 |
| Glioma IV | 17 (59%) | 19 (31%) | .024 |
| **Error rate** |  |  |  |
| Overall ER (%) | 8.43 ± 4.92 | 4.37 ± 2.93 | <0.001 |
| Left ER (%) | 8.87 ± 4.66 | 4.55 ± 3.06 | <0.001 |
| Right ER (%) | 6.71 ± 4.93 | 5.01 ± 3.67 | 0.232 |
| Semantic ER (%) | 2.23 ± 2.11 | 0.97 ± 1.12 | <0.001 |
| Motor ER (%) | 6.20 ± 4.54 | 2.59 ± 1.64 | <0.001 |

Values shown are mean±SD or n (proportion). ER = Error rate (amount of positive TMS stimulations divided by the sum of positive and negative TMS stimulations).

### 2.2 Data Acquisition

#### 2.2.1 MRI

MRI data were acquired using a Siemens 3T Skyra system (Erlangen, Germany) at Charit University Hospital, Berlin, Department of Neuroradiology. T1 weighted images were acquired with the use of the 3D magnetization-prepared rapid gradient echo (MPRAGE) sequence with TR/TE/TI 2300/2.32/900 ms flip angle = 8°, field of view (FOV) = 230 × 230 mm^2^, matrix size 256 × 256, 192 sagittal slices, 0.9mm isotropic resolution.(Mugler and Brookeman, 1990)

#### 2.2.2 rTMS language mapping

Navigated rTMS language mapping was performed with nTMS eXimia NBS version 3.2.2, Nexstim NBS 4.3 and NexSpeech module (Nexstim Oy, Helsinki, Finland). The rTMS language mapping was conducted as previously reported.(Schwarzer *et al*., 2018) The degree of discomfort or pain during the mapping was evaluated with visual analogue scale (VAS). Not properly named images, or not immediately named or named with difficulties were excluded from the following rTMS examination to limit misnaming unrelated to rTMS. The baseline was performed up to three times, in case errors were still made in the second baseline. The images remaining after the final baseline cycle were used for rTMS mapping, which covered the perisylvian cortex of both hemispheres.(Schwarzer *et al*., 2018) The rTMS coordinates were exported as text files for subsequent analysis and spatial normalization. rTMS induced language errors were classified into two categories: Performance errors (e.g. stuttering), phonological errors (vocalization), no responses (absence of reply) and hesitation errors (delayed response) were grouped in category I, semantic errors (misnaming error) were grouped in category II. This grouping allowed to disentangle errors that could have been caused by issues with uttering a response alone (Category I) from errors that pointed directly to an impairment of semantic processing (Category II) in picture naming.

#### 2.2.3 Aphasia grading

The severity of aphasia was assessed preoperative using the Berlin Aphasia Score (BAS). The BAS is used and developed by physicians of the Charit University Hospital and adapted to the Aachener Aphasia Test.(Huber *et al*., 1980) The test classifies patients into 4 categories: 0 = no aphasia, 1 = mild aphasia, 2 = moderate aphasia, 3 = severe aphasia, A = motor accented, B = sensory accented.(Picht *et al*., 2013; Schwarzer *et al*., 2018) All patients with a BAS score of 0 were grouped into the non-aphasic cohort, others were classified as aphasic patients.

#### 2.2.4 Spatial normalization & anatomical labelling

In order to optimize the registration process to MNI ICM 152 space, we skull-stripped all T1 MPRAGE data sets applying the ANTs brain extraction tool in combination with the public ANTs/ANTsR IXI brain template (https://doi.org/10.6084/m9.figshare.915436.v2) prior to MNI space registration.(Avants *et al*., 2011) Furthermore, semi-automated lesion segmentations were generated with ITK-Snap and provided as binarized masks (Fig. 1).(Yushkevich *et al*., 2006) All patients’ T1 MPRAGE image data sets were registered to the normalized space MNI ICBM 152 non-linear 6th Generation Symmetric Average Brain Stereotaxic Registration Model using the Advanced Normalization Tools (ANTs) software with the Symmetric Normalization (SyN) transformation model.(Grabner *et al*., 2006; Avants *et al*., 2011) The registration matrix files were used to subsequently register the T1 MPRAGE data sets based rTMS coordinates to MNI ICM 152 space as well. The rTMS coordinates were mapped to the Automated Anatomical Labeling (AAL) parcellation to define their anatomical volumes of interest (AVOI).(Tzourio-Mazoyer *et al*., 2002) The AAL labelling offers a whole brain parcellation of 90 AVOIs, excluding the cerebellum (116 in total).(Tzourio-Mazoyer *et al*., 2002) The AAL coordinates were gathered using SPM12 (http://www.fil.ion.ucl.ac.uk/spm) and SPM-based viewing program xjView (http://www.alivelearn.net/xjview8/).(Ashburner and Friston, 1999) Category I & II errors were counted for each AVOI. After merging all rTMS spots, we calculated the error rate (ER) in each AVOI. The ER was calculated by dividing the number of rTMS stimulations with error responses by the number of total rTMS stimulations.

**Fig 1.**
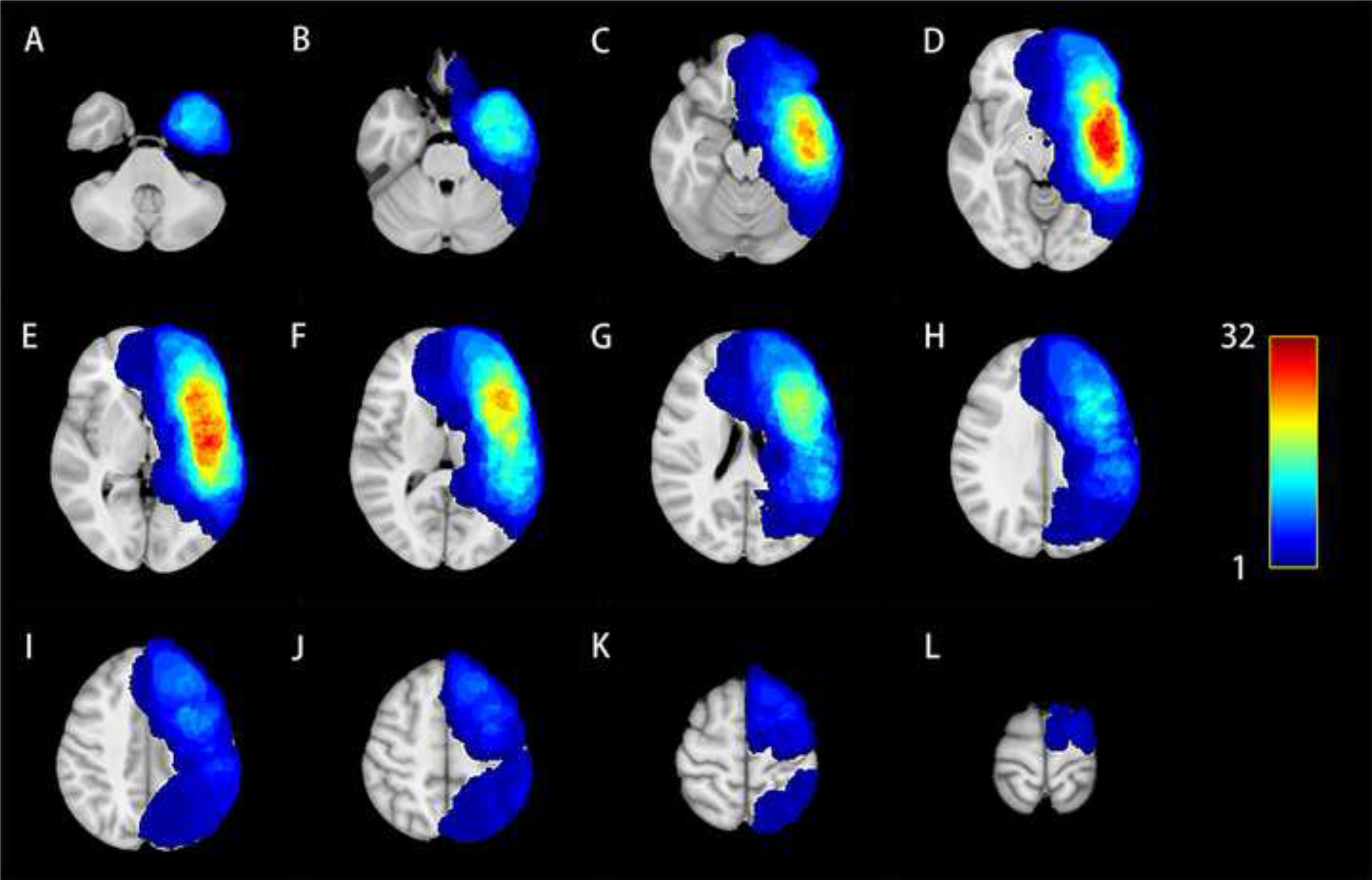
Lesion map. The figure shows axial maps in MNI space (z axis = -40 – 70) with an increment of 10mm. All lesions are located in the perisylvian area. The regions of maximal overlap (red) are in the peri-sylvian area.

### 2.3 SVM

With the progression of computational power over the last two decades, SVM has received growing attention and is increasingly being used in biomedicine. SVMs are supervised machine learning models and aim to classify data points by maximizing the margin between classes.(Cortes and Vapnik, 1995)

SVM can be used for nonlinear classification using kernel tricks by implicitly mapping inputs to high-dimensional feature spaces using various kernel functions.(Liu *et al*., 2019) In the present study we used the linear kernel function, followed by the recursive feature elimination (RFE) method for feature selection using the training data set to classify the patients into aphasic or non-aphasic groups. Features which had less than 10 % missing values were included to fit in the SVM model. Missing data were interpolated with the mean or median, depending on whether the variables followed normal distribution or not. All 90 patients were selected for training and subsequent testing. A grid search strategy was used to tune the classifier parameters. A leave-one-out cross validation (LOO-CV) approach was applied for training. Parameter C was determined to be 1.15. Due to the unbalanced data, we adjusted the weight of the aphasia group to 2.0. For machine learning coding we used MATLAB R2014b (MathWorks, Natick, MA, US) with LIBSVM.(Chang and Lin, 2011) We evaluated the performance of the SVM model by sensitivity, specificity, its overall accuracy as well as area under the receiver operating characteristic curve (AUC). The overall accuracy is the ratio of correctly predicted classification to the entire cohort in the aphasic or non-aphasic group.

### 2.4 Statistical analysis

Statistical analysis was performed by using MATLAB R2014b (MathWorks, Natick, MA, US) and SPSS22 IBM SPSS, Armonk, New York, US). To compare continuous variables, two tailed Student’s *t*-tests or Mann–Whitney U tests were performed separately, according to normally distributed data. Significant effects were considered at P = 0.05. Fisher’s exact test (for expected values less than five) or Pearson’s chi-squared test (larger values) were applied for the comparison of parameter variables.

### 2.5 Data availability

The data that support the findings of this study are not publicly available due to information that could compromise the privacy of the research participants but are available from the corresponding author on reasonable request.

## 3. Results

### 3.1 Patients

29 (32.2%) of the recruited 90 patients presented with presurgical aphasic language disorders. The demographic data and comparison between aphasic and non-aphasic patients are provided in Table 1. There were no significant differences in sex or tumor size in relation to aphasia (*P* = .272, χ2 = 1.207; *P* = .982, *t* = .023, respectively). Aphasic patients were older (56.69±13.64) than non-aphasic patients (45.13±12.84), with a highly significant difference of *P* < .001. The tumor locations were distributed into frontal lobe (34), temporal lobe (39), parietal lobe (10) and insular lobe (7).

### 3.2 Presurgical rTMS mapping

Presurgical rTMS speech mapping was successful in all patients and generally well tolerated. 22 patients were only mapped on their left hemisphere due to fatigue or decreasing attention level. The mapping results of frontopolar and temporopolar cortices were not considered for analysis due to the discomfort evoked by the rTMS mapping in these areas (Fig. 2). The mean VAS score during rTMS mapping was 3.9±2.9 in the left and 3.7±2.8 in the right hemisphere. The ER of the entire brain mapping in the aphasia group (7.49 [IQR 5.82,9.53]) was significantly higher than the ER (3.48 [IQR 2.48,5.79]) of the entire brain mapping in the non-aphasia group (*P* < .001, Z = -4.606, η^2^ = .238).

**Fig 2.**
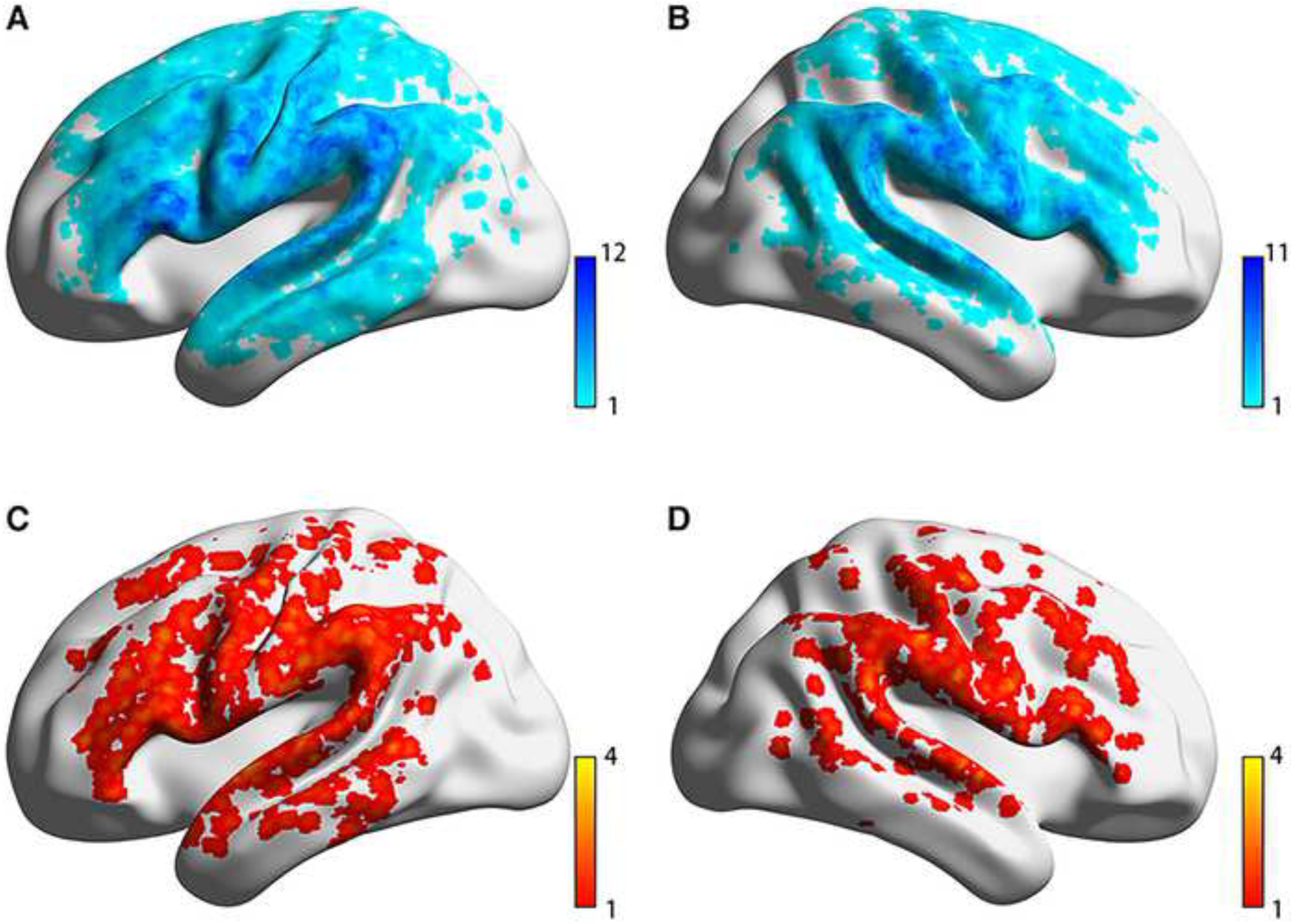
Voxel-wise TMS stimulation spot distribution of all patients in MNI space: (A) Left negative spots. (B) Right negative spots. (C) Left positive spots. (D) Right positive spots. The numbers indicate the count of TMS positive or negative stimulations per voxel.

The aphasic patients showed a significantly higher ER in their left hemisphere (aphasia group: 8.00 [IQR 6.32,10.80] compared to the non-aphasic patients (3.79 [IQR 2.48,6.47]; *P* < .001, Z = -4.718, η^2^ = .250) and a non-significant difference of ER in their right hemisphere (aphasia group: 5.38 [IQR 2.73,10.28]; non-aphasia group: 3.65 [IQR 2.33,6.70]; *P* = .232, Z = - 1.187, η^2^ = .022). In relation to category I ERs (performance errors, phonological errors, no response and hesitation errors), the aphasia group showed higher ERs than the non-aphasia group (aphasia group: 5.54 [IQR 2.83,8.35]; non-aphasia group: 2.19 [IQR 1.40,3.57]; *P* < .001, Z = - 4.753, η^2^ = .254). The same observation was made for category II ERs (semantic errors), the aphasia group showed higher ERs than the non-aphasia group (aphasia group: 1.79 (IQR 0.59,3.43); non-aphasia group: 0.65 (IQR 0,1.61); *P* = .003, Z = -2.959, η^2^ = .098). The overall rTMS positive spot distribution showed no clear pattern, not favoring specific cortical areas (Fig. 2). Moreover, the closely matching cortical distribution of both positive and negative rTMS spot distribution further demonstrates the non-occurrence of a particular pattern.

### 3.3 AAL labelling

The analysis of aphasic and non-aphasic patients in relation to rTMS ERs distribution showed specific cortical patterns. The AVOIs of ERs were calculated for each patient (Supplementary Table 1). To increase the reliability of the ER results, AVOIs which were stimulated less than 250 times in the sum of all patients, resulting in a total inclusion of 28 AVOIs (Fig. 3 & Table 2).

Overall 11 AVOIs showed significant differences between the aphasia group and non-aphasia group (Table 2), revealing a perisylvian pattern: Frontal_Inf_Tri_L (*P* = .012, Z = -2.56, η^2^ = .084), Frontal_Inf_Tri_R (*P* = .036, Z = -2.11, η^2^ = 0.081), Frontal_Mid_L (*P* = .025 Z = - 2.25, η^2^ = .072), Parietal_Inf_L (*P* = .002, Z = -3.24, η^2^ = 0.133), Parietal_Inf_R (*P* = .049, Z = - 1.98, η^2^ = .067), Precentral_L (*P* < .012, Z =-2.51, η^2^ = .078), Rolandic_Oper_L (*P* = .020, Z= -2.34, η^2^ = .070), Rolandic_Oper_R (*P* = .026, Z = -2.23, η^2^ = .082), SupraMarginal_L (*P* < .001, Z = -3.54, η^2^ = .161), Temporal_Mid_L (*P* = .008, Z = -2.67, η^2^ = 0.089) and Temporal_Sup_L (*P* = .048, Z = -1.98, η^2^ = .046).

**Table 2.** ER distribution.

| Region, Side (L/R) | Overall ER (%)<br>MED (IQR) | | *P* | *Z-score* | $\eta^2$ | Category I ER (%)<br>MED (IQR) | | *P* | *Z-score* | $\eta^2$ | Category II ER (%)<br>MED (IQR) | | *P* | *Z-score* | $\eta^2$ |
| --- | --- | --- | --- | --- | --- | --- | --- | --- | --- | --- | --- | --- | --- | --- | --- |
|  | Non-aphasic<br>patients | Aphasic<br>patients |  |  |  | Non-aphasic<br>patients | Aphasic<br>patients |  |  |  | Non-aphasic<br>patients | Aphasic<br>patients |  |  |  |
| Angular_L | 0 (0) | 0 (11.11) | .237 | -1.182 | .023 | 0 (0) | 0 (8.33) | .465 | -0.731 | .009 | 0 (0) | 0 (0) | .721 | -0.355 | .002 |
| Angular_R | 0 (3.33) | 0 (11.65) | .322 | -0.989 | .020 | 0 (0) | 0 (8.13) | .455 | -0.748 | .012 | 0 (0) | 0 (0) | .636 | -0.474 | .005 |
| Frontal_Inf_Oper_L | 0 (9.02) | 3.85 (15.38) | .343 | -0.949 | .012 | 0 (7.69) | 0 (12.64) | .717 | -0.362 | .002 | 0 (0) | 0 (0) | .334 | -0.966 | .012 |
| Frontal_Inf_Oper_R | 0 (10.00) | 0 (12.50) | .815 | -0.234 | .001 | 0 (6.08) | 0 (9.77) | .692 | -0.396 | .003 | 0 (0) | 0 (0) | .496 | -0.682 | .008 |
| Frontal_Inf_Tri_L | 2.95 (7.69) | 8.33 (12.48) | .012 | -2.557 | .084 | 2.34 (6.42) | 6.90 (12.5) | .066 | -1.839 | .043 | 0 (0) | 0 (4.51) | .062 | -1.869 | .045 |
| Frontal_Inf_Tri_R | 0 (5.26) | 5.88 (14.05) | .036 | -2.111 | .081 | 0 (5.20) | 0 (7.74) | .306 | -1.023 | .019 | 0 (0) | 0 (5.73) | .046 | -1.992 | .071 |
| Frontal_Mid_L | 0 (6.93) | 6.25 (14.22) | .025 | -2.252 | .072 | 0 (5.07) | 0 (7.89) | .516 | -0.650 | .006 | 0 (0) | 0 (5.26) | .009 | -2.611 | .098 |
| Frontal_Mid_R | 0 (9.52) | 4.45 (11.88) | .545 | -0.606 | .006 | 0 (5.56) | 4.26 (9.88) | .283 | -1.074 | .020 | 0 (0) | 0 (0) | .713 | -0.367 | .002 |
| Frontal_Sup_L | 0 (1.32) | 0 (25.60) | .129 | -1.520 | .054 | 0 (0.99) | 0 (10.71) | .729 | -0.347 | .003 | 0 (0) | 0 (16.07) | .004 | -2.873 | .192 |
| Frontal_Sup_R | 0 (0) | 0 (10.56) | .179 | -1.343 | .046 | 0 (0) | 0 (5.56) | .125 | -1.533 | .060 | 0 (0) | 0 (0) | .975 | -0.032 | .000 |
| Parietal_Inf_L | 0 (6.56) | 6.67 (10.56) | .002 | -3.243 | .133 | 0 (4.55) | 4 (11.76) | .030 | -2.176 | .060 | 0 (0) | 0 (5.26) | .085 | -1.725 | .038 |
| Parietal_Inf_R | 0 (4.95) | 2.94 (16.07) | .049 | -1.981 | .066 | 0 (4.00) | 0 (9.09) | .391 | -0.858 | .012 | 0 (0) | 0 (0) | .099 | -1.650 | .046 |
| Parietal_Sup_L | 0 (13.84) | 0 (0) | .287 | -1.078 | .025 | 0 (8.38) | 0 (0) | .518 | -0.647 | .009 | 0 (0) | 0 (0) | .157 | -1.414 | .043 |
| Parietal_Sup_R | 0 (7.69) | 0 (16.07) | .976 | -0.304 | .002 | 0 (0) | 0 (0) | .588 | -0.542 | .007 | 0 (0) | 0 (0) | .345 | -0.944 | .020 |
| Postcentral_L | 3.71 (6.90) | 4.08 (11.76) | .357 | -0.922 | .010 | 2.60 (5.26) | 3.85 (8.90) | .144 | -1.460 | .026 | 0 (2.49) | 0 (0) | .833 | -0.211 | .001 |
| Postcentral_R | 4.35 (7.77) | 5.56 (11.15) | .445 | -0.763 | .009 | 4 (6.30) | 1.72 (5.56) | .566 | -0.573 | .005 | 0 (0) | 0 (3.34) | .217 | -1.235 | .024 |
| Precentral_L | 2.47 (6.67) | 8.02 (12.50) | .012 | -2.511 | .078 | 1.95 (6.40) | 4.21 (10.28) | .267 | -1.109 | .015 | 0 (0) | 0 (4.08) | .002 | -3.168 | .124 |
| Precentral_R | 0 (6.17) | 4.67 (14.84) | .233 | -1.194 | .023 | 0 (5.73) | 0 (8.17) | .926 | -0.093 | .000 | 0 (0) | 0 (5.30) | .015 | -2.443 | .095 |
| Rolandic_Oper_L | 0 (0) | 0 (25.00) | .020 | -2.341 | .070 | 0 (0) | 0 (20.83) | .030 | -2.180 | .061 | 0 (0) | 0 (0) | .296 | -1.044 | .014 |
| Rolandic_Oper_R | 0 (0) | 6.25 (16.07) | .026 | -2.231 | .082 | 0 (0) | 0 (10.82) | .147 | -1.452 | .035 | 0 (0) | 0 (5.36) | .195 | -1.295 | .028 |
| SupraMarginal_L | 4.55 (8.33) | 9.55 (10.98) | .001 | -3.541 | .161 | 1.32 (6.78) | 8.01 (11.11) | .008 | -2.644 | .090 | 0 (0) | 0 (7.55) | .075 | -1.778 | .040 |
| SupraMarginal_R | 5.26 (10.67) | 4.27 (10.26) | .379 | -0.881 | .012 | 4.35 (10) | 1.43 (5.24) | .179 | -1.345 | .029 | 0 (0) | 0 (5.61) | .329 | -0.976 | .015 |
| Temporal_Mid_L | 2.67 (8.00) | 8.33 (9.88) | .008 | -2.672 | .089 | 0 (5.42) | 5.88 (9.23) | .026 | -2.238 | .063 | 0 (1.19) | 0 (3.65) | .284 | -1.071 | .014 |
| Temporal_Mid_R | 0 (9.09) | 0 (2.50) | .817 | -0.231 | .001 | 0 (5.13) | 0 (5.61) | .818 | -0.230 | .001 | 0 (0) | 0 (2.63) | .286 | -1.067 | .025 |
| Temporal_Pole_Sup_L | 0 (1.61) | 0 (10) | .748 | -0.321 | .002 | 0 (0) | 0 (0) | .919 | -0.102 | .000 | 0 (0) | 0 (0) | .875 | -0.158 | .000 |
| Temporal_Pole_Sup_R | 0 (6.25) | 0(3.13) | .948 | -0.065 | .000 | 0 (0) | 0 (0) | .443 | -0.767 | .012 | 0 (0) | 0 (3.13) | .071 | -1.806 | .067 |
| Temporal_Sup_L | 3.18 (9.55) | 7.69 (12.50) | .048 | -1.979 | .046 | 0 (5.91) | 4.54 (8.27) | .109 | -1.602 | .030 | 0 (0) | 0 (4.76) | .026 | -2.240 | .058 |
| Temporal_Sup_R | 1.56 (0) | 5.90 (10.83) | .229 | -1.202 | .023 | 0 (5.44) | 4.29 (7.85) | .254 | -1.141 | .021 | 0 (0) | 0 (0) | .892 | -0.135 | .000 |

Values shown are median (IQR). Asterisks denote significant differences between non-aphasic and aphasic patients at $p < 0.05$ . $\eta^2$ represents the effect size of Mann-Whitney U test: $\eta^2 = Z^2 / (n - 1)$ .

**Fig 3.**
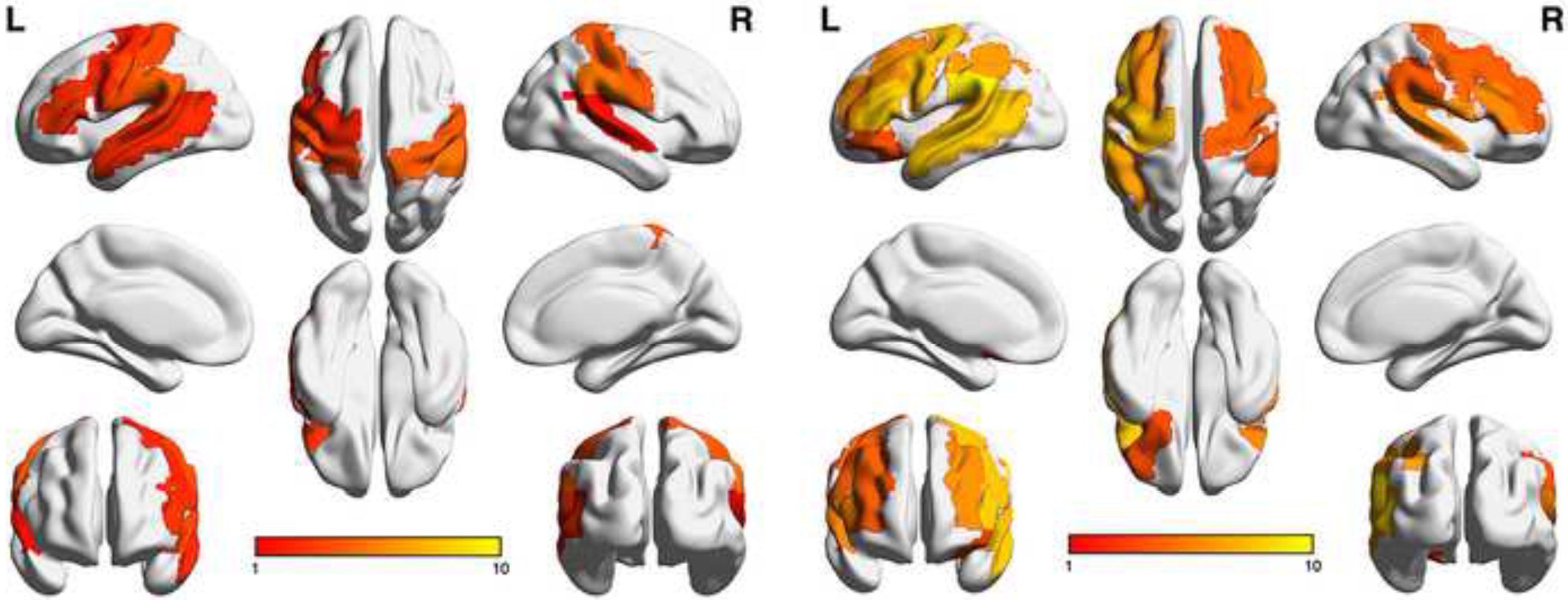
Visualization of overall ER distribution in relation to AAL parcellation in non-aphasic (left) and aphasic (right) groups.

Furthermore, the subgroup analysis of category I (performance errors, phonological errors and hesitation errors) ERs showed distinct cortical left Rolandic and temporal and right frontal patterns. Here, 4 AVOIs showed significant differences between the aphasia group (higher ERs) and non-aphasia group within category I (Table 2): Parietal_Inf_L (*P* = .030, Z = - 2.18, η^2^ = .060), Rolandic_Oper_L (*P* = .030, Z =- 2.18, η^2^ = .061), SupraMarginal_L (*P* <.008, Z = -2.64, η^2^ = .090), Temporal_Mid_L (*P* = .026, Z = -2.24, η^2^ = .063).

The subgroup analysis of category II (semantic errors) ERs showed as well a different distinct left frontal and right central cortical pattern (Table 2). 6 AVOIs showed significant differences between the aphasia group and non-aphasia group, including following AAL AVOIs: Frontal_Inf_Tri_R (*P* = .046, Z = -1.99, η^2^ = .071), Frontal_Sup_L (*P* = .004, Z = -2.87, η^2^ = .192), Frontal_Mid_L (*P* = .009, Z = -2.61, η^2^ = .098), Precentral_L (*P* = .002, Z = -3.17, η^2^ = .124), Precentral_R (*P* = .015, Z = -2.44, η^2^ = .095), Temporal_Sup_L (*P* = .026, Z = -2.24, η^2^ = .058).

### 3.4 SVM classification

We built three different SVM models. One based purely on the ERs, one with a feature for age and another with a feature for tumor location. By adding a feature for tumor location, the model could not be improved compared to the pure ER model. However, by adding a feature for age, the model was improved (improvement of accuracy from 75.6% to 81.1%). The above 11 ER AVOIs with significant differences were taken as input features for the SVM-RFE model. After RFE was embedded within a LOO-CV framework, three regions were selected as the most important features. Fig. 4 and Table 3 illustrate the weight of these three regions (Frontal_Inf_Tri_R, SupraMarginal_L, Parietal_Inf_L). For the test, sensitivity was 89.7%, specificity was 82.0%, overall accuracy was 81.1% and AUC was 88.7%. Fig. 5 illustrates the model’s receiver operating characteristic (ROC) curve and indicates the AUC.

**Table 3.** Weights learned with SVM classification with LibSVM, with linear kernel C = 1.15.

| Feature | Weight |
| --- | --- |
| Frontal_Inf_Tri_R | 2.06 |
| SupraMarginal_L | 2.05 |
| Parietal_Inf_L | 1.80 |
| Age | 2.95 |

**Fig 4.**
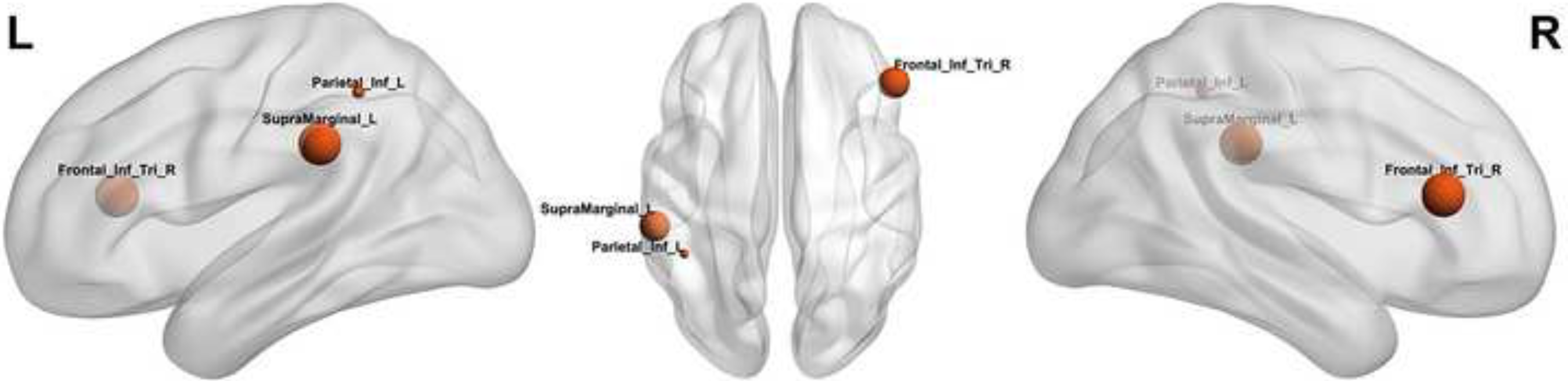
Visualization of AVOIS-based SVM weights. The visualization illustrates the different weights of AVOIS classified by the SVM model, shown in left sagittal, dorsal and right sagittal views.

**Fig 5.**
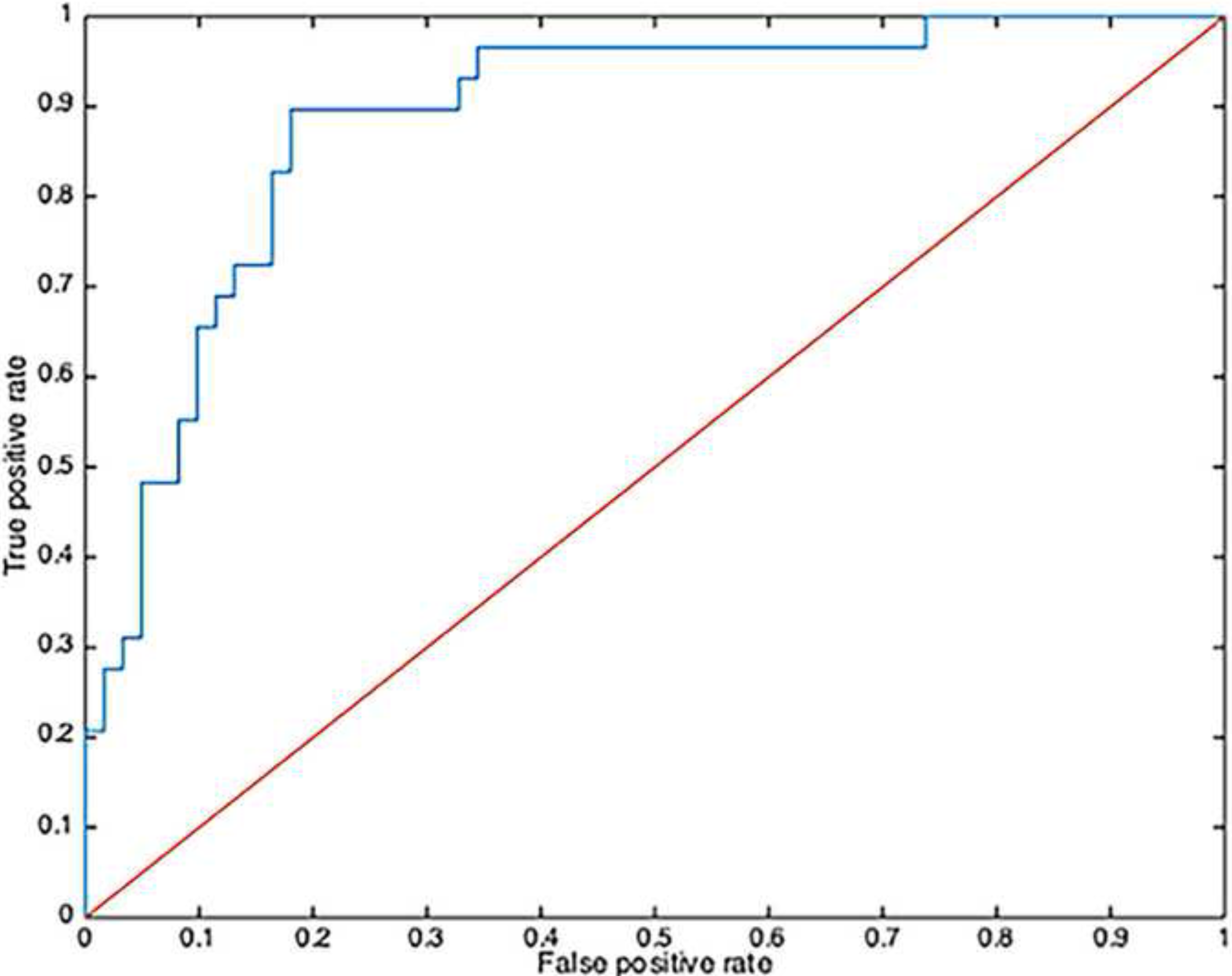
ROC for the SVM model with AUC = 88.7%.

## 4. Discussion

In this study, we examined the language network in a spatially normalized cohort of 90 rTMS mapped glioma patients applying group analysis methodology. Our results confirm current neuroplasticity models from stroke research with regard to aphasia as well as tumor-induced language neuroplasticity findings.(Thiel *et al*., 2005; Piai, 2019; Stefaniak *et al*., 2019) Interestingly, the pattern of overall positive rTMS spots does not clearly differ from the pattern of overall negative rTMS spots (Fig 1). Furthermore, the rTMS-based analysis does not confirm traditional localizationist models of the language network, but rather illustrates a perisylvian distribution of cortical areas where the patients’ language network was disturbed by rTMS. This observation is in line with other studies, illustrating a pivotal role of left and right frontal, precentral, central and parietal areas in general (Table 2).(Stefaniak *et al*., 2019)

The connectomic configuration of the language network is confirmed by the left and right distribution of category I & II errors, showing a left hemispheric frontal and perisylvian distribution as well as presenting right frontal and precentral error susceptibility to rTMS. A recent study focused on the results of MEG in patients with tumors in the language area of the dominant hemisphere where semantic properties shifted to the right hemisphere.(Piai, 2019) fMRI experiments show asymmetric left lateralization patterns for language function in healthy subjects and thus weaken a possible hypothesis of an a priori bihemispheric configuration.(Raemaekers *et al*., 2018)

In contrast to the rTMS ERs analysis, the SVM results do not show a proportional increase of ER in all areas, but rather an increment in specific right frontal and left parietal areas, such as Frontal_Inf_Tri_R, SupraMarginal_L, Parietal_Inf_L. This result directs us towards an interpretation of a functional shift, indicating a functional relevant involvement of the right frontal area in relation to aphasia. Furthermore, Table 2 indicates that patients with aphasia show an increased rTMS error susceptibility left and right frontally as well as a larger right perisylvian distribution of increased ERs. Moreover, while it is of crucial importance to interpret these results with regard to the tumor location, the location feature didn’t affect the SVM model, whereas the age feature improved the SVM model. This result is confirmed by an early study predicting language dysfunction, that correlated age and tumor grade but not tumor location with aphasia.(Recht *et al*., 1989) This finding supports the notion that general tumor induced network disconnection is relevant to aphasia and not necessarily related to specific lesion locations. Additionally, the results emphasize the decreasing potential for neuroplasticity with age, confirming earlier studies.(Lu *et al*., 2004)

The relationship of aphasia and the right hemisphere’s susceptibility to rTMS induced errors points towards rTMS being a marker for language network impairments and resulting reorganization. By analyzing the overall increase of errors in aphasic patients it can be stated that even after baseline correction of the object-naming image stack, patients with neuropsychological impairment tend to make more errors during mapping of both hemispheres. This unspecific effect can be attributed to purely false positive errors or to an unspecific increase of the language network’s susceptibility to rTMS disruption of language processing. In a previous study a cut-off value of 28% errors during baseline naming of the object naming task was identified, above which the naming task cannot be reliably enough performed by the subjects.(Schwarzer *et al*., 2018) Yet, the SVM results demonstrate that the overall increase in ER over both hemispheres in the aphasic patients manifests in a disproportional increase of errors on the right cortex, sustaining a specific effect of left hemispheric pathology on functional reorganization, specifically highlighting the relevance of rIFG, as previously claimed.(Thiel *et al*., 2005; Rosler *et al*., 2014) The functional relevance of the right hemispheric areas with increased vulnerability towards rTMS is still unclear. In stroke studies recruitment of the rIFG after left hemispheric stroke was associated with unfavorable outcome compared to cases with predominant regional ipsilateral recruitment.(Winhuisen *et al*., 2007) Yet, in brain tumor patients, there is evidence that a shift towards the right hemisphere is associated with better outcome after left hemispheric surgery, sustaining the hypotheses of increased functional reserve in patients with more bilateral distribution of language function.(Ille *et al*., 2016)

### 4.1 Limitations

The number of rTMS stimulations per area is heterogenous, with the discomfort evoked by the current rTMS methodology limiting cortical coverage. In addition, the use of only one task (object naming) can induce a systematic error due to potential location specificity.(Krieg *et al*., 2017) Also, error annotation is user dependent leading to difficulties to compare results across institutions and warranting objective measures for analysis of language performance. Finally, as long as task performance is necessary, cognitive mapping in general is heavily depending on the patient’s performance, inducing a difficult to control bias.

## 5. Conclusion

The results of this study based on group analysis, cortical parceling and machine learning classification with an SVM model, show that the pattern of rTMS-induced errors in aphasic patients differs distinctly from the pattern in non-aphasic patients, with the right inferior frontal gyrus in particular indicating increased vulnerability to rTMS. Furthermore, the use of SVM classification offers new perspectives for rTMS data analysis and allows new interpretations of the method, which contributes to our understanding of rTMS in tumor patients with challenged neuronal networks. While reliable mapping of the functional language-network remains a major challenge in individual brain tumor patients, the results of this study suggest that machine learning adds to detecting distinct patterns of functional reorganization in patients with language eloquent brain tumors. This study constitutes the first machine learning based classification of rTMS language mapping results.

## Acknowledgements

The brain networks shown in Fig. 2, 3 & 4 were visualized with the BrainNet Viewer (http://www.nitrc.org/projects/bnv/).(Xia *et al*., 2013)

^1^

**Abbreviations**

AAL: automated anatomical labeling
AAT: Aachener aphasia test
AUC: area under receiver operating characteristic curve
AVOI: anatomical volumes of interest
BAS: Berlin aphasia score
ER: error rate
IQR: interquartile range
MNI: Montreal neurological institute
RFE: recursive feature elimination
ROC: receiver operating characteristic
rTMS: Repetitive transcranial magnetic stimulation
SVM: support vector machine
TMS: transcranial magnetic stimulation
VAS: visual analogue scale

## Credit Author Statement

All authors did the research and wrote this manuscript. They have made a direct and substantial contribution to the study reported and participated to a sufficient degree. Z. Wang participated in the data collection. Z. Wang, L. Fekonja and T. Picht conceived and designed the study. Z. Wang, L. Fekonja and F. Dreyer analyzed and interpreted the data and did the statistical analyses. All authors participated in writing the article and provided a critical discussion and conclusions.

